# Automated visual acuity estimation by optokinetic nystagmus using a stepped sweep stimulus

**DOI:** 10.1101/2024.02.19.23300472

**Authors:** Jason Turuwhenua, Zaw LinTun, Mohammad Norouzifard, Misty Edmonds, Rebecca Findlay, Joanna Black, Benjamin Thompson

## Abstract

**Purpose:** Measuring visual acuity (VA) can be challenging in adults with cognitive impairment and young children. We developed an automatic system for measuring VA using Optokinetic Nystagmus (OKN).

**Methods:** VA-OKN and VA by ETDRS (VA-ETDRS) were measured monocularly in healthy participants (n=23, age 30±12). VA was classified as reduced (n=22, >0.2 logMAR) or not (n=24, ≤0.2 logMAR) in each eye. VA-OKN stimulus was an array of drifting (5 deg/sec) vanishing disks presented in descending/ascending size order (0.0 to 1.0 logMAR in 0.1 logMAR steps). The stimulus was stepped every 2 seconds, and 10 sweeps were shown per eye. Eye tracking data determined when OKN activity ceased (descending sweep) or began (ascending sweep) to give an automated sweep VA. Sweep traces were randomized and assessed by a reviewer blinded to VA-ETDRS. A final per sweep VA and VA-OKN was thereby determined.

**Results:** A single randomly selected eye was used for analysis. *<u>VA deficit group:</u>* There was no significant difference between overall mean VA-OKN and VA-ETDRS (p>0.05, paired t-test) and the r^2^ statistic was 0.84. The 95% limits of agreement were 0.19 logMAR. *<u>No VA deficit group:</u>* There was a 0.24 logMAR bias between VA-OKN and VA-ETDRS and no correlation was found (r^2^ = 0.06). However, the overall sensitivity/specificity for classification was 100%.

**Conclusions:** A robust correlation between VA-ETDRS and VA-OKN was found. The method correctly detected a VA deficit.

**Translational relevance:** OKN is a promising method for measuring VA in cognitively impaired adults and pre-verbal children.

## INTRODUCTION

Visual acuity (VA) is the quantitative measure of the ability to discriminate fine detail. It is arguably the single-most important functional measure of vision, and it is used to diagnose and monitor a range of visual disorders including refractive error, macular degeneration and amblyopia.^1, 2^ However, measuring VA is challenging in people who may lack the cognitive ability to perform standard VA tests, such as adults with neurological impairment or very young children.^3, 4^

There are two objective approaches available to estimate VA in such patients: 1) visual evoked potentials (VEPs) or 2) optokinetic nystagmus (OKN).^5^ VEPs are electrical responses measured from the visual cortex that arise in response to structured targets shown to the observer. The stimulus is typically constructed from stripes, gratings or checkboards, which are typically swept in steps from low to high frequency with cortical activity measured at each step. VA is determined by regression applied to higher spatial frequency activities in order to find the x-axis crossing of the activity versus time graph,^6^ the idea being that cortical activity decreases as the stimulus’ spatial frequency approaches threshold. However, despite the fact that the sweep VEP is a rapid and well understood method^7, 8^ in clinical practice it requires a trained operator and expensive/complex equipment which makes it difficult to implement.^5, 9^

OKN is a reflexive oculomotor response of the eye consisting of stereotyped slow tracking (the slow phase or SP) and quick, opposite direction, re-fixation movements (the quick phase or QP) that occur in response to a drifting pattern.^10^ OKN manifests as a highly distinctive “beating” of the eye, which appears in eye tracking data as a repeated “sawtooth” pattern on a displacement versus time plot as shown in **Fig.1**. In clinical practice, OKN is elicited by the induction method: a vertically striped drum is rotated in front of the patient. If the stimulus is visible to the observer it will elicit the involuntary OKN response whilst if it is invisible to the patient then it will not. Varying the spatial frequency of the stimulus (by altering the distance of the drum to the eye), gives an estimate of VA as the minimum grating size that elicited OKN. However, the spinning drum technique is a highly subjective procedure.

**FIGURE 1.**
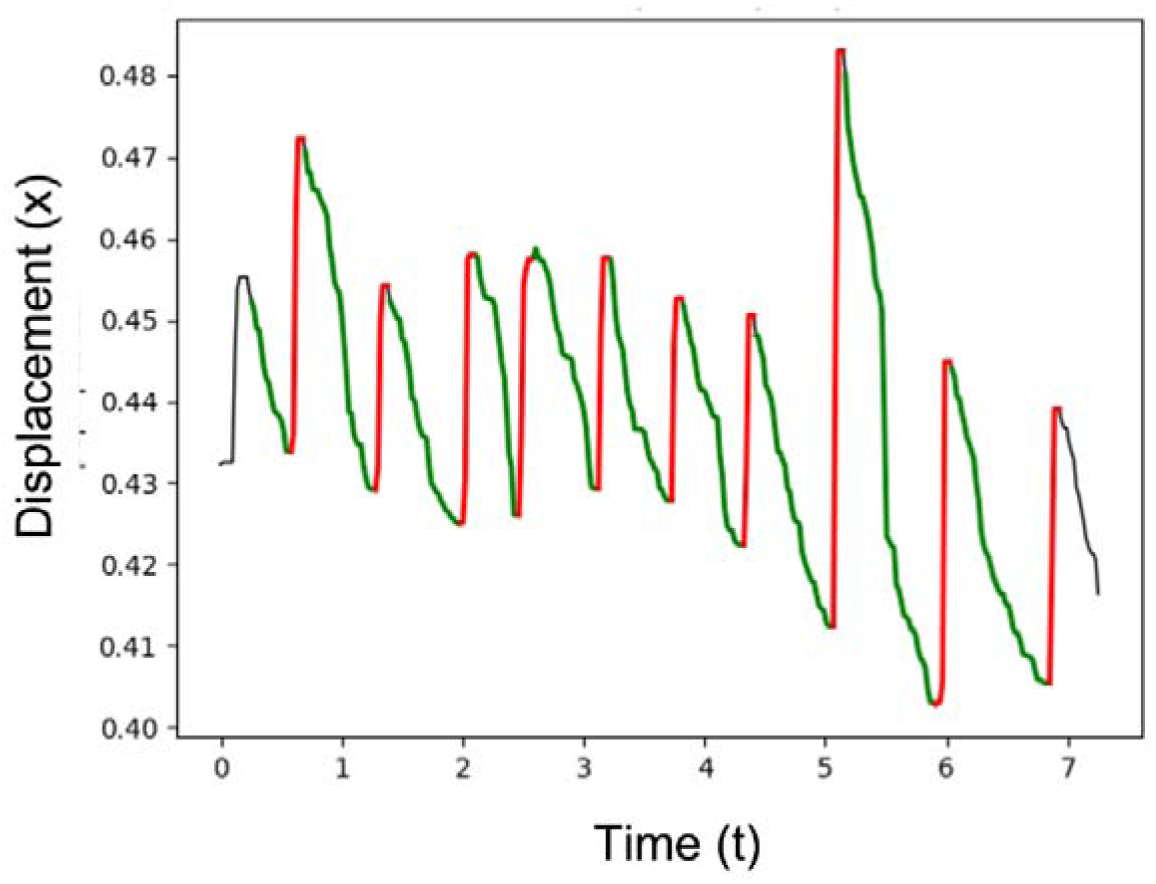
Optokinetic nystagmus. Automatically detected slow phases are shown in green, and quick phases are shown in red.

The general concept of objective estimation of VA using OKN is long established,^11–13^ yet its application as an objective probe of visual function is only now receiving renewed interest.^14–19^ Hyon et. al. incorporated video-oculography and used a high contrast (85%) striped stimulus drifting at 10 deg/sec on a display to test distance vision.^20^ The induction method was used to measure a minimum stripe size. The suppression method,^5^ in which a fixation target of varying visibility is used to halt the response, was also used to determine the minimum dot size needed to stop the optokinetic response. In that work, r^2^ statistics of 0.57 and 0.83 were reported against logMAR VA using induction/suppression paradigms respectively. Shin et al. ^21^ used a similar experimental paradigm applied to a range of disease states. The reported correlation coefficients corresponded to r^2^ statistics between 0.38-0.88 (induction) and 0.006-0.76 (suppression). However, Aleci et al. noted that the precision and accuracy in these previous studies was low,^14, 22^ and that the stripe stimulus velocity imposed severe restrictions on the range of acuities that could be tested. Aleci et al. therefore proposed a slowly moving (1.43 and 2.86 deg/sec) and low contrast (20%) row of optotypes. Using an induction paradigm an r^2^ statistic of 0.74 with VA (measured by Sloan optotypes) was reported for co-operative participants, noting also the inclusion of a noise component added to the symbols to lower the visibility of larger optotypes. Using the same paradigm, Aleci et. al ^22^ subsequently reported r^2^ statistics of 0.63 and 0.70 with subjective VA for AMD and Cataract patients respectively.

Our own approach shares a similar motivation to that of Aleci et al. Firstly, we avoid a striped stimulus. Instead we propose a novel stimulus consisting of an array of slow drifting (5 deg/sec) vanishing disk optotypes. As shown in **Fig. 2(a)**, each disk of the array comprises a brighter central disk (mean contrast 87%) which determines the angular size of the stimulus calibrated in logMAR units. Crucially, the disk has a darker surround (mean contrast −14%) that averages out the central disk at threshold in accordance with the vanishing principle.^23, 24^ In doing so, we aim to mitigate the velocity and width restrictions imposed by stripes, avoid very low contrast (20%) stimuli that have been used previously, and remove the need for additional noise. Moreover, an advantage of the logMAR specification is that it avoids reliance on arbitrary units used previously.^11, 20–22^

**FIGURE 2.**
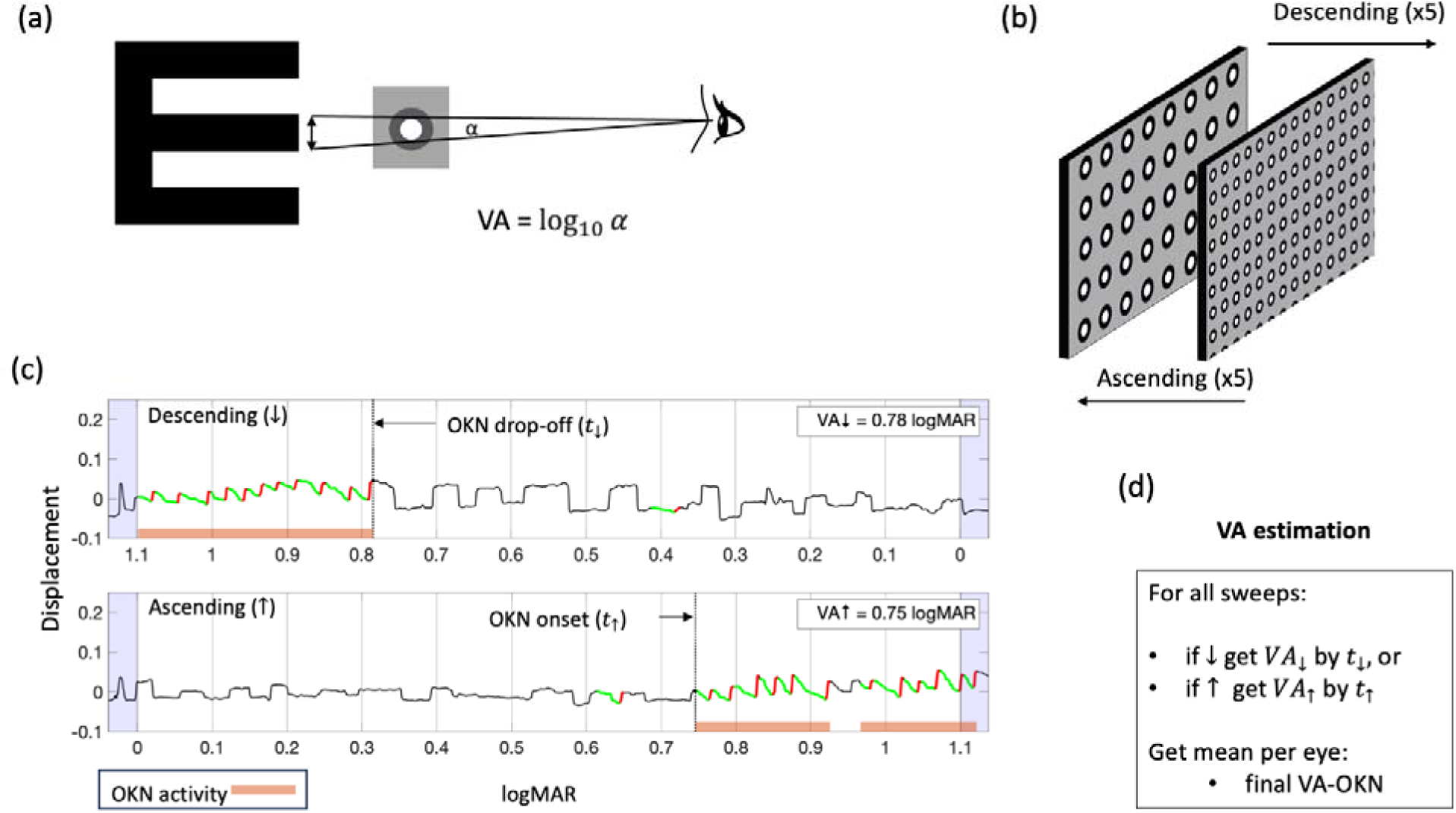
(a) The VA for the vanishing disk stimulus is determined by the angular size (α) of the central brighter disk. (b) The stimulus is a horizontally drifting array of vanishing disks, which is swept in descending/ascending order. (c) Analysis of eye tracking data. Top panel shows results for a descending sweep whilst bottom panel shows results for an ascending sweep. The OKN activity is shown in orange along the bottom. The OKN drop-off point/onset point determines a VA for a sweep. (d) Final VA-OKN is the average of a total of 10 sweeps.

In this work we use repeating stepped ascending and descending sweeps varying the logMAR size of the stimulus at each step as shown in **Fig. 2(b)**. Aleci et al. used a similar adaptive sweep that was subjectively driven by an observer. However we use eye tracking to objectively measure eye displacement data during each sweep, which is processed by an OKN detection system. **Fig. 2(c)** shows eye tracker data for a descending (top panel) and ascending sweep (lower panel). The figure shows OKN activity for the sweep (shown in orange) which is used to determine the OKN drop-off and onset times (the times when OKN stops or starts depending on sweep direction). For a given descending sweep the drop-off point is the last time point of OKN activity, whilst for the ascending sweep the onset time point is the first point of OKN activity. Finally we perform VA scoring as shown in **Fig. 2(d)**. We propose a *per* sweep VA based on standard letter-by-letter scoring used for VA charts.^25^ Our key modification is to replace letter counting with the drop-off/onset times at which OKN disappears/appears to provide a VA. An overall final VA is found by averaging over all sweeps.

We provide proof-of-concept of the overall approach in a small cohort of people with and without a visual deficit. We determine a VA by automation, and then obtain a final VA-OKN after assessment by a reviewer blinded to VA by ETDRS (VA-ETDRS). Final VA-OKN is compared to VA-ETDRS measured using the Electronic Visual Acuity (EVA) tester (Emmes, USA). Potential improvements to the presented work are provided in the discussion.

## DESCRIPTION OF PROCESSING PIPELINE

### VA scoring for OKN

Our scoring is based on the Early Treatment of Diabetic Retinopathy Study (ETDRS) chart. The chart consists of 5 letters per line. Each line differs by 0.1 logMAR, and each letter has a value of 0.02 logMAR. In a standard letter-by-letter scoring method^25^ the patient reads out aloud each letter from the largest logMAR line (*VA_max_*) to the smallest (*VA_min_*). *VA_↓_* is then given by,

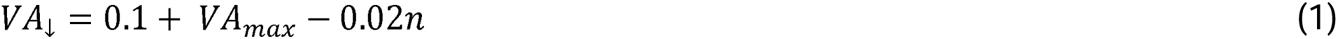

where *n* is the number of correctly identified letters, and the arrow indicates the direction of reading (from largest to smallest letters). It is also possible to move from the smallest letters of size *VA_min_* to the largest. In this case we have,

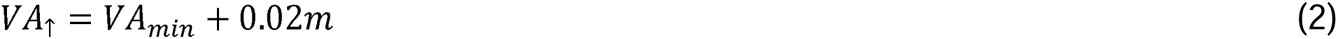

where *m* is now the number of incorrectly counted letters, and the up arrow indicates the direction of reading (from smallest to largest letters).

Crucially, our approach proposes to replace letters in Eqns. (1) and (2) by the *time* at which the OKN response to stimulus of decreasing/increasing size either *stops*, *t*_↓_ or commences *t*_↑_ (descending and ascending sweeps respectively). Eqn (1) now becomes

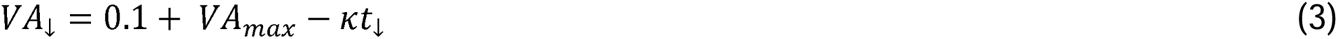

for the descending sweep, and Eqn. (2) becomes

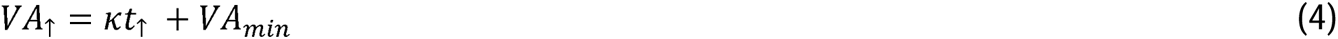

for the ascending sweep. Here *k* is the absolute step-rate in logMAR/sec. It is noted here that using the earliest/latest time to replace letter counting is a convenient heuristic that we test empirically. The methods for determining *t*_↓_ and *t*_↑_ that we implemented will be described further below.

If a sweep did not generate detectable OKN then the VA was set to the upper limit VA_max_ +0.1 on the understanding that it would be greater than or equal to this result. This could be expected if a participant did not see the stimulus, or did not generate detectable OKN during the test, or if there was an OKN detection/measurement failure.

### OKN detection

Objective OKN detection was facilitated, in this work, by an already published method. The reader is encouraged to consult that work for further details.^26^ In brief, this approach segmented the signal in the incoming eye tracking signal into SP-QP pairs. The SP was found by looking for a ramping displacement in the known stimulus direction. The QP was then found as a quick re-setting in the opposite direction subsequent to the SP. Candidate OKN was thresholded based on criteria such as amplitude, duration and velocity^26^ to eliminate unlikely detections. At this point, if remaining OKN was found to be repeating (2 or more consecutive detections) it was considered to be *valid* OKN. The “repeating OKN” criterion has proven to be a strong feature in work to date and also appears in the work of others.^18, 26, 27^ We also detected groups of at least three unconnected instances of OKN that fell within a sliding 2-second window. We observed that this situation could arise if OKN was present but was missed by the detector (e.g., low amplitude OKN). This “intermittent” OKN rule was used to mitigate cases where the “repeating OKN” criterion was not activated. Any OKN points found by these criteria were labelled as *valid* OKN.

### OKN activity & VA estimation

The two empirical criteria of the previous section together give an instantaneous *OKN activity*,

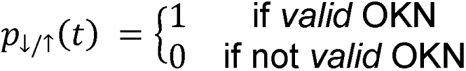

where the arrows specify an descending or ascending sweep respectively. Given a descending sweep, we define the OKN drop-off point *t*_↓_ as the point at which OKN ceases. More formally this is

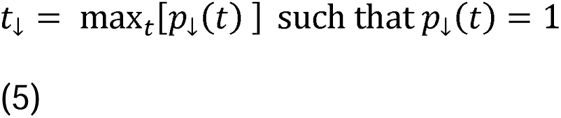

which is the *maximum* time point *t*_↓_ of *OKN activity*. If no drop-off time is found then *VA*_↓_ ≥ 0.1 + *VA_max_* as determined by substituting *t*_↓_=0 in Eqn (3). Given an ascending sweep, the OKN onset time *t*_↑_ occurs at the *minimum* time of OKN activity,

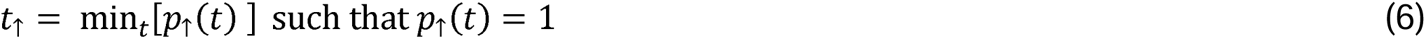

If this point does not exist then we set *t*_↑_ to the time corresponding to the end of the last presentation, and VA will also be *VA*_↑_ ≥ 0.1 + *VA_max_*. In any case, given any otherwise found *t*_↑_ or *t*_↓_ value, we enter the result of Eqn. (5) into Eqn. (3) for *VA*_↓_, or Eqn. (6) into Eqn (4) for *VA*_↑_.

**Fig 2(c)** demonstrates this process in the case of two subsequent sweeps (descending/ascending respectively). The green/red regions indicate all instances of *potential* OKN. Our process identifies these as *valid* OKN (or not) to yield the OKN activity. The OKN activity is shown in orange along the base of each sweep (presence corresponds to activity). The drop-off and onset-points are the times of last activity (top panel) and first activity (bottom panel) respectively as marked as dotted lines. These are then transformed into logMAR VAs using Eqns. (3) and (4), and are reported in the top right corner of each panel.

## METHODS

### Participants

The research followed the tenets of the Declaration of Helsinki. The study was approved by the University of Auckland Human Participants Ethics Committee (UAHPEC20318). Participants (n=11) with reduced VA due to refractive error were recruited, as were participants with no refractive error (n=12). Both eyes of the participants were measured monocularly without refractive correction with occlusion of the non-tested eye.

### VA-ETDRS

Participants’ VAs were measured using the EVA system in single-letter presentation mode. Letters were flanked by crowding bars. Testing distance was 3m. VA-ETDRS was the average of up to 5 repeated measurements taken consecutively within a single measurement session. Participants with a logMAR VA-ETDRS of ≤0.2 logMAR were classified as “no VA deficit”. Those with a VA of >0.2 logMAR were classified as belonging to a “reduced VA” group.^29^ These groups were analysed separately.

### VA-OKN

#### Equipment

The viewing distance was 3 meters. The stimulus was presented on a 27-inch monitor (Dell S2716DG). This monitor had the nVidia Ultra-Low Motion Blur (ULMB) function engaged to minimize the presence of motion blur inherent to light-emitting diode displays.^28^ The refresh rate was 85 Hz. Eye tracking data was measured using Pupil Invisible glasses (Pupil Labs, Berlin, Germany) with sampling at 200Hz and down-sampling to 50Hz. The stimulus was written using the jsPsych framework for behavioural experiments, and the experiment was presented using the Chrome web-browser window running in full-screen mode. The system was co-ordinated by a standard desktop PC using a combination of Javascript and Python. Further offline analysis of this data after data collection was performed using Matlab (Natick, VA).

#### Stepped sweep stimulus

The average contrast (with respect to the background) was 87% (central disk) and −14% (annulus). The angular size of the central disk was the central disk size in logMAR units. A sweep stepped through 11 logMAR levels from 0.0 to 1.0 in 0.1 logMAR steps in descending/ascending order. The stimulus stepped every 2 seconds (step rate was 0.05 logMAR/sec) and stimulus drift speed was 5 deg/sec for the entirety of a sweep. The total presentation time for a single sweep was 22 seconds. The first/last presentations were extended before the start and end of the measurement interval by 0.75 sec (giving a total presentation time of 23.5 seconds) to allow some time for OKN to either initiate before or pass beyond the analysis interval. These appear as the blue regions on either side of a sweep presented in **Fig 2(b)** and were not included for analysis. As mentioned already, the stimulus was shown as 5 descending/ascending interleaved sweeps (a total of 10 sweeps). The time to test an eye was about 4 minutes.

#### VA estimation

An automated per sweep VA (*VA*_↑_ or *VA*_↓_) was determined using the objective analysis described in the previous section. OKN detection was performed using a single “best set” of OKN detection parameters that were found empirically and applied across all data collected.

The total number of sweeps processed was 460 (23 participants * 10 sweeps* 2 eyes = 460). Automated VA-OKN was averaged over the 10 sweeps per eye. Sweep data was reviewed at the time of data-collection. Measurement of an eye could be repeated if unwanted artefacts (e.g., due to excessive blinking, or noisy signal) were identified.

After this stage of analysis, a further assessment step was performed by a reviewer blinded to VA-ETDRS. Measured sweeps were randomized and resulting sweep data traces (for example as shown in **Fig 2(c)**) were provided to the reviewer. The reviewer was asked to identify their own estimate of a final per sweep VA starting with the automated sweep results. The average of these reviewed sweep VAs provided a final VA-OKN.

## RESULTS

### Results for final VA-OKN

A summary of results is presented in **Table 1** and **Table 2** for the “reduced VA” group and “no VA deficit” groups respectively (n=23 participants and a total of 46 eyes). Results below are reported for a randomly selected eye, noting that the right eyes for P-02 and P-04 were purposefully excluded from analysis because their final VA-OKN results were unbounded.

**TABLE 1.**
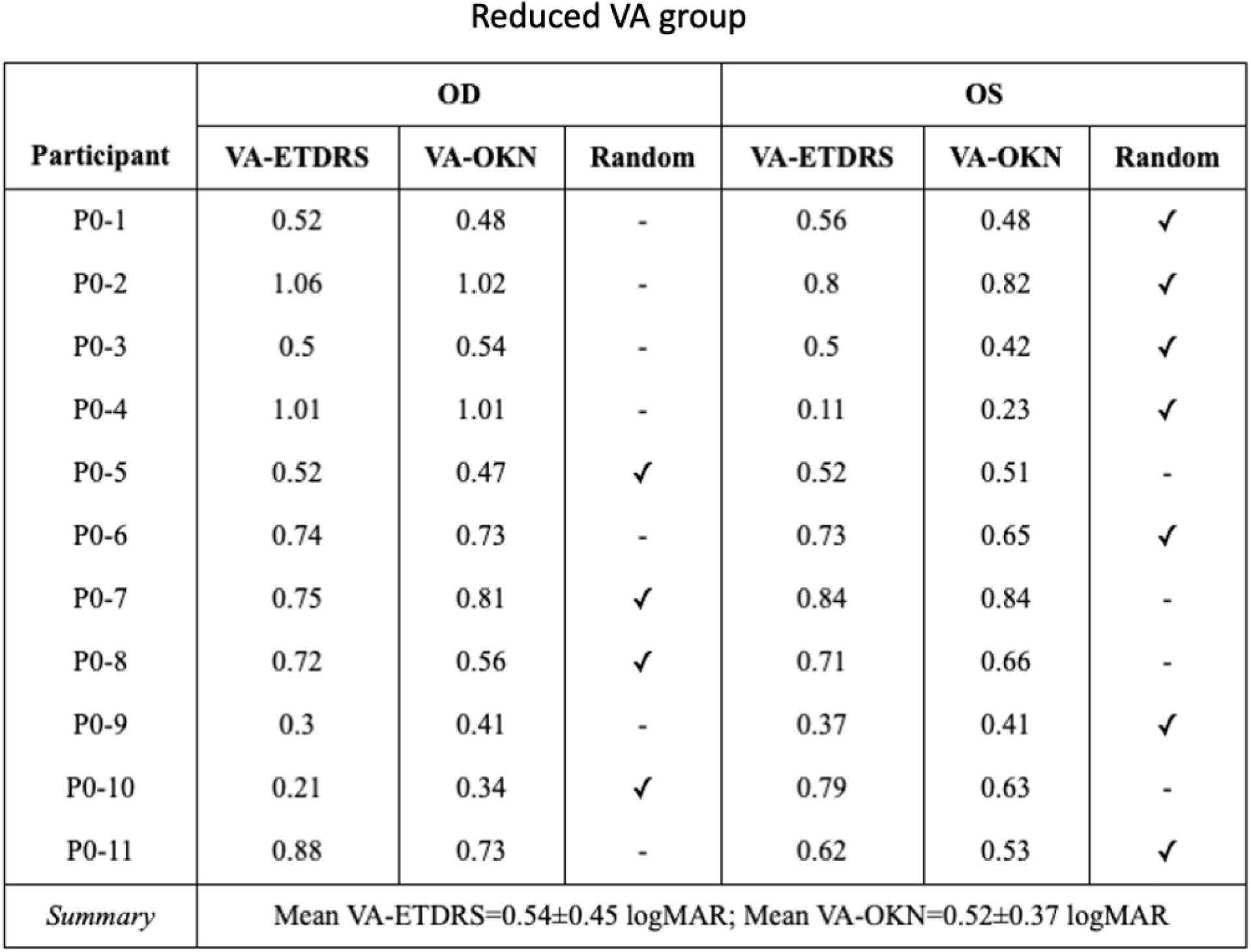
VA-ETDRS and VA-OKN for the reduced VA cohort. Data is shown for both eyes, but only a randomly selected eye was used for reported comparisons (indicated by a tick).

**TABLE 2.** VA-ETDRS and VA-OKN for the no VA deficit cohort. Data is shown for both eyes, but only a randomly selected eye was used for reported comparisons (indicated by a tick).

| Participant | OD |  |  | OS |  |  |
| --- | --- | --- | --- | --- | --- | --- |
|  | VA-ETDRS | VA-OKN | Random | VA-ETDRS | VA-OKN | Random |
| P1-12 | -0.26 | 0.03 | ✓ | -0.24 | 0.02 | - |
| P1-13 | -0.21 | 0 | ✓ | -0.14 | 0.03 | - |
| P1-14 | -0.18 | 0.01 | ✓ | -0.11 | 0 | - |
| P1-15 | -0.16 | 0.12 | - | -0.22 | 0.12 | ✓ |
| P1-16 | -0.12 | 0.05 | - | -0.16 | 0.06 | ✓ |
| P1-17 | -0.05 | 0.07 | ✓ | -0.07 | 0.01 | - |
| P1-18 | -0.14 | 0.05 | - | -0.19 | 0.13 | ✓ |
| P1-19 | -0.14 | 0.02 | - | -0.19 | 0.01 | ✓ |
| P1-20 | -0.1 | 0.03 | ✓ | 0 | 0.02 | - |
| P1-21 | -0.22 | 0 | - | -0.22 | 0.01 | ✓ |
| P1-22 | -0.3 | 0.02 | ✓ | -0.3 | 0.02 | - |
| P1-23 | -0.15 | 0.13 | ✓ | -0.16 | 0.04 | - |
| *Summary* | Mean VA-ETDRS=-0.19±0.13 logMAR; Mean VA-OKN=0.05±0.1 logMAR |  |  |  |  |  |

#### Reduced VA group

VA-OKN was 0.52±0.37 logMAR (mean ± 2SD). VA-ETDRS was 0.54±0.45 logMAR. There was no significant difference between the two measures (p=0.55 >0.05, t=-0.61, paired t-test). **Fig 3(a)** presents a scatter plot comparing VA-OKN and VA-ETDRS for the “deficit” group only. The r-squared value (r^2^) was 0.84. The corresponding Bland-Altman diagram in **Fig 3(b)** plots the differences between VA-OKN and VA-ETDRS against their means. The limits of agreement (LoA) were 0.19 logMAR and the mean difference was −0.02 logMAR. The mean difference was not significantly different from zero by the result comparing VA-OKN and VA-ETDRS above.

**FIGURE 3.**
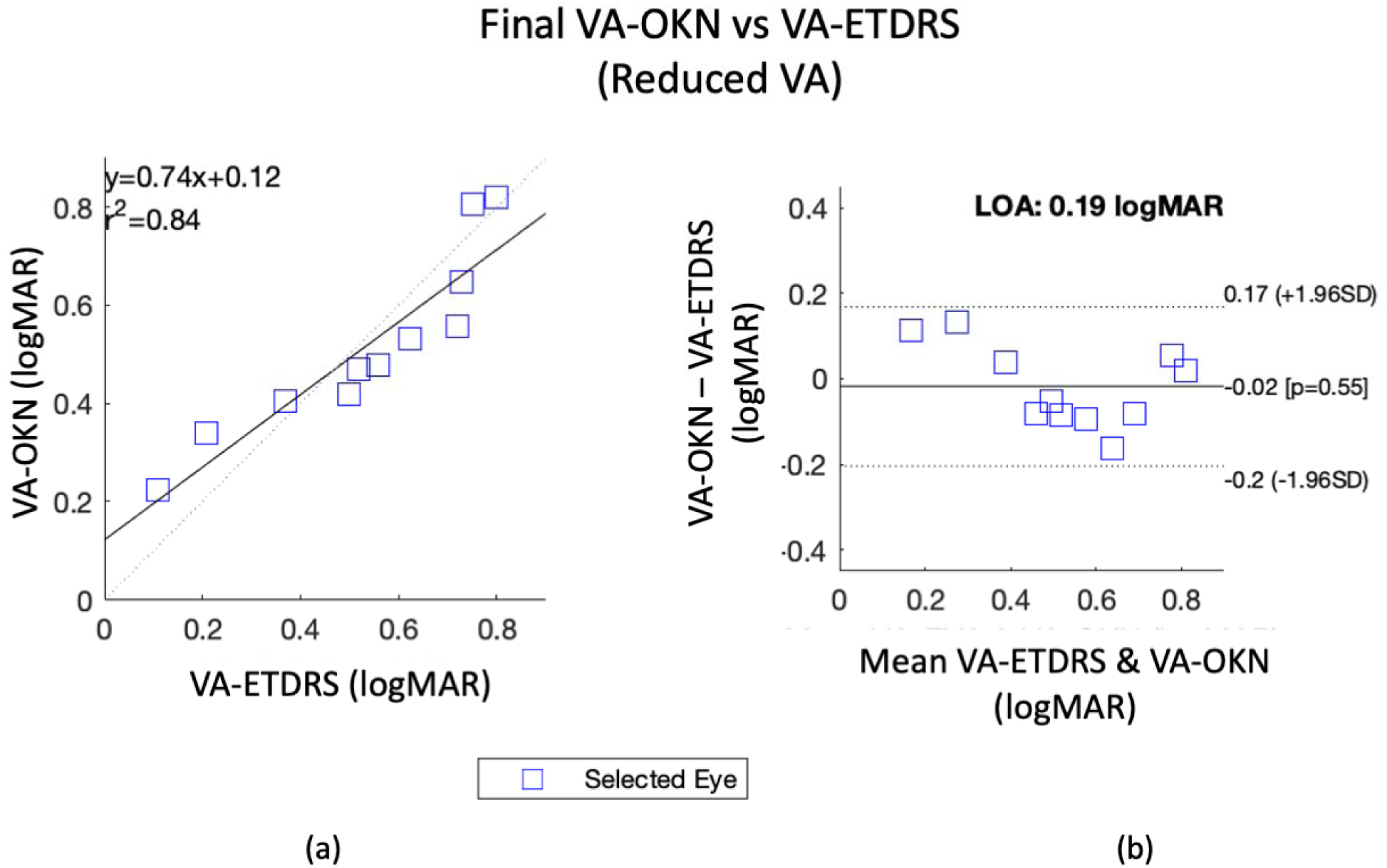
(a) The correlation plot for the reduced VA group. The regression line is a solid line, the ideal line is dotted. (b) Bland-Altman diagram shows difference between VA-OKN and VA-ETDRS against their means.

#### No VA deficit group

VA-OKN was 0.05 ± 0.10 logMAR. VA-ETDRS was −0.19 ± 0.13 logMAR. There was a significant difference between the two groups (p=0.00 <0.05, t=8.50, paired t-test). This value was also lower/better than the floor of the OKN measurement device (0.0 to 1.1 logMAR). **Fig 4(a)** presents a scatter plot that directly compares VA-ETDRS and VA-OKN directly against each other for the “no deficit” group. The r^2^ statistic was 0.06 indicating no correlation between the two VA measures. The Bland-Altman diagram in **Fig 4(b)** produced an LoA of 0.14 logMAR and mean difference of 0.24 logMAR. The mean difference was significant as already determined by the result comparing VA-OKN to VA-ETDRS.

**FIGURE 4.**
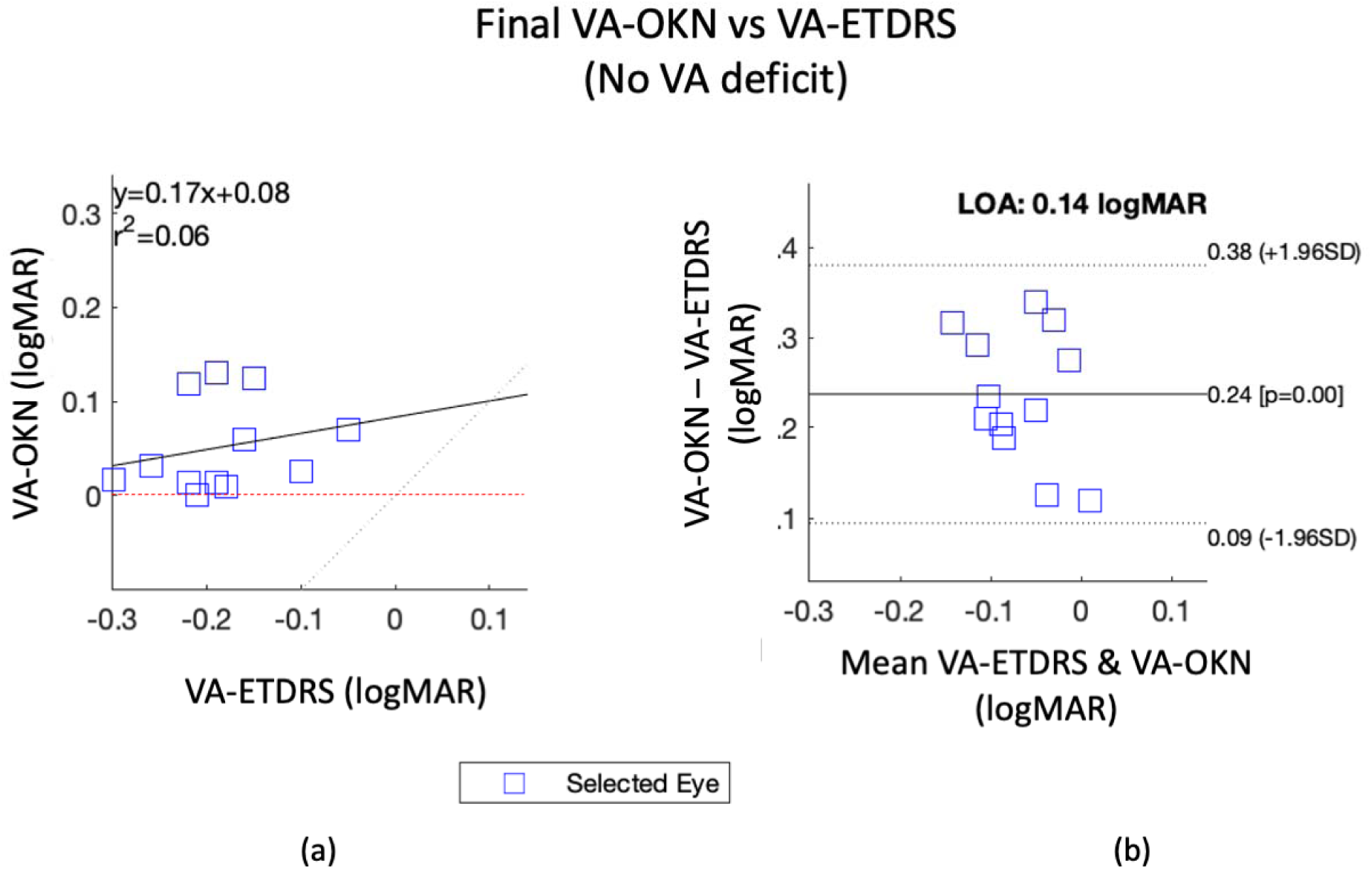
(a) The correlation plot for the no VA deficit group. The regression line is a solid line, the ideal line is dotted. Red line shows the floor of the OKN device (0.0 logMAR). (b) Bland-Altman diagram shows difference between VA-OKN and VA-ETDRS against their means.

#### Screening performance

There were no instances in which VA-OKN misclassified the presence/absence of VA deficit as determined by VA-ETDRS. Overall the sensitivity/specificity for detecting a VA deficit was 100%.

### The effect of an expert review of automated VA-OKN

#### Reduced VA group

The overall mean for the deficit group found by automation was 0.54 ± 0.36 logMAR. This was not changed significantly by an expert review of the results (p=0.80>0.05, t=3.67, paired t-test). The r^2^ for final VA-OKN vs VA-ETDRS was 0.02 higher than the automated result alone. The LoA for final VA-OKN vs VA-ETDRS was 0.1 logMAR lower than the automated result.

#### No VA deficit group

The no VA deficit group mean was 0.07±0.10 logMAR by automation. The automated VA-OKN result was not changed significantly by the manual review (p=0.21>0.05, t=-1.32, paired t-test). The r^2^ and LoAs were unchanged.

#### Screening performance

The pre-review sensitivity was 92%. The specificity was 100%. The reduced sensitivity occurred because a single eye was incorrectly classified as having VA deficit (VA-OKN=0.21>0.2 logMAR) by automation.

## DISCUSSION

In this paper we presented an automated pipeline for measurement of VA by OKN. The final VA-OKN results were encouraging. For those in the “VA deficit” group, for which the range of VA-ETDRS results fell within the measurement range of the VA-OKN measurement system, we observed a strong correlation (r^2^=0.84) between VA-OKN and VA-ETDRS. This r^2^ value compares favourably to results reported previously for VA by OKN between 0.53-0.83.^14, 22, 30^

Previous work has been somewhat limited by the use of arbitrary units,^11, 14, 20, 21^ which have constrained comparisons between OKN-induction stimuli spatial characteristics and standard letter based VA. In this work, Bland-Altman analysis indicated a degree of agreement between VA-OKN and VA-ETDRS. The LoAs for the VA deficit group were 0.19 logMAR. These limits are comparable to results from VEP based VA assessment noting two studies by Bach et. al. ^7, 31^ which indicated an accuracy of about 0.3 logMAR. Although not a direct comparison, the results are on the order of the inherent (test-retest) variability found for ETDRS charts of 0.11 to 0.25.^29, 32^ Whilst the present results are encouraging, we also recognize as with VEP,^33^ that OKN based testing is not the same as recognition VA measured with a chart. The exploration of these differences in healthy participants and those with ocular disease would characterize the method more fully and would be the subject of further work.

For the “no VA deficit” group we found a significant bias of 0.24 logMAR between VA-OKN and VA-ETDRS. VA-ETDRS was significantly better/lower than VA-OKN. Whilst the lowest/best VA-OKN that could be measured by our device was 0.0 logMAR, it is likely that there was also an inherent bias due to the stimulus itself. Hamilton et. al.^8^ reported similar biases between sweep VEP and recognition VA of 0.03 to 0.3 log units for normally sighted participants.

Extending the range of the stimulus to provide direct comparison in the lower/better logMAR range is a possibility for future work. A larger cohort with a wider spread of VA-ETDRS values should provide a clearer picture of performance overall.

VA-OKN accurately classified all participants as “no VA deficit” or “reduced VA” using a screening threshold of 0.2 logMAR. This result suggests that VA-OKN may provide a viable screening option, particularly for adults and children who are unable to engage with current vision screening techniques, which frequently involve measurement of visual acuity using symbols or letters.

It should be noted that the automated pipeline *did* require operator supervision. The expert review detected a misclassification error, and improved r^2^ and LoA overall. We observed that OKN detection could fail because of excessive blinking, false detections caused by noisy signal or incorrect classification of saccades as OKN; none of which were handled directly by the present method. Fortunately, we had the opportunity to address such issues by re-measuring our co-operative adult participants. Our system was not tested on adults with cognitive impairment or young children in this work, and we recognize that the opportunity to remediate measurement problems may not be available to the operator in such subjects. Improvements could be made to defend against unwanted artefacts (*e.g.*, by pausing the experiment in the presence of excessive blinking), or to improve the robustness of data acquisition/analysis (*e.g.*, by explicit detection and removal of purely saccadic movements). It may be that we could skip a given logMAR level if it was robustly detected (descending sweep) or, simply stop the sweep altogether similarly (ascending sweep) when OKN commenced. Recently, work has been done in improving the classification of OKN by machine learning methods^35, 36^ which we expect will have significant benefits for such efforts described here. For now, we can only emphasize the need for supervision and caution when interpreting results produced by the system.

Finally, we note that the parameters of the sweep such as stimulus velocity, step duration, and specific contrasts used in the construction of the stimulus were chosen largely empirically. Although the resulting stimulus presented here appeared to be effective, there remains a large parameter space for the OKN induction stimulus, and the optimum combination of spatial and temporal parameters is still being established.

## CONCLUSION

We have presented a clinically inspired method for determining VA threshold using an automated response. The results support the concept of objective assessment of VA using OKN. Overall, the method here aims to progress toward clinically applicable objective estimation of VA using the optokinetic response. Results to date are encouraging, but further investigation into OKN based VA is warranted.

A simplified demonstration of the sweeping vanishing disk stimulus is available at: bit.ly/4ayJhwq

## Data Availability

All data produced in the present work are contained in the manuscript

## ACKNOWLEDGEMENT

Support is acknowledged from the Health Research Council of New Zealand (HRC21/514).

## Notes

**Financial support:** Health Research Council of New Zealand (HRC 21/514). BT is supported by InnoHK and the Hong Kong Government.

### Competing Interest Statement

The authors have declared no competing interest.

### Funding Statement

The study was supported by the Health Research Council of New Zealand (HRC 21/514). BT is supported by InnoHK and the Hong Kong Government.

### Author Declarations

The study was approved by the University of Auckland Human Participants Ethics Committee. The ethics approval reference is UAHPEC20318

